# The extent and impact of vaccine status miscategorisation on covid-19 vaccine efficacy studies

**DOI:** 10.1101/2024.03.09.24304015

**Authors:** Martin Neil, Norman Fenton, Scott McLachlan

## Abstract

It is recognised that many observational studies and randomised control trials reporting high efficacy for Covid-19 vaccines suffer from various biases. Systematic review identified thirty-seven studies that suffered from one particular and serious form of bias called miscategorisation bias, whereby study participants who have been vaccinated are categorised as unvaccinated up to and until some arbitrarily defined time after vaccination occurred. Simulation demonstrates that this miscategorisation bias artificially boosts vaccine efficacy and infection rates even when a vaccine has zero or negative efficacy. Furthermore, simulation demonstrates that repeated boosters, given every few months, are needed to maintain this misleading impression of efficacy. Given this, any claims of Covid-19 vaccine efficacy based on these studies are likely to be a statistical illusion or are exaggerated.

## 1. Introduction

Considerable attention has been given to the reported high efficacy for the Covid-19 vaccines in observational studies and randomised control trials, and how many of these studies have exhibited signs of measurement biases and confounding (Reeder, 2021, Fung, Jones & Doshi, 2023; Heying & Weinstein, 2023; Ioannidis, 2022; Fenton & Neil, 2023, Lataster 2024). One major kind of bias takes the form of miscategorisation, whereby study participants who have been vaccinated are miscategorised as unvaccinated up to and until some arbitrarily defined time after vaccination occurred (typically up to 14 or 21 days). This bias, which has been seen to take several different types, all of which help exaggerate vaccine efficacy, has recently become known colloquially as the ‘cheap trick’ (Fenton & Neil, 2023).

To identify the different types of miscategorisation bias and evaluate how widespread it is, we conducted a review of Covid-19 observational vaccine studies to identify those studies that have employed miscategorisation bias and we have simulated the effects of this bias on measures of vaccine efficacy.

This review reveals that, up to February 2024, 37 observational studies and randomised control trials on Covid-19 vaccines have employed different types of this bias, with variants including straightforward miscategorisation from one category to another, miscategorising the vaccinated as having unverified vaccination status, uncontrolled reporting of vaccination status and excluding those vaccinated from the study. Many of the studies have applied one or more of these biases within time periods from one week to three.

Our simulation model demonstrates that this bias artificially boosts vaccine efficacy in all cases, and with the application of repeated ‘booster’ vaccinations, the efficacy of repeated Covid-19 vaccines could be maintained at artificially high levels in perpetuity. Furthermore, in tandem with this the infection rate would likewise be artificially elevated and would be higher for the unvaccinated cohort compared to the vaccinated cohort, further compounding misleading claims that a Covid-19 vaccine reduces infection rates when it does not.

The paper is structured as follows: In Section 2 we review the work on biases in Covid-19 vaccine studies. In Section 3 we describe the search method by which relevant studies were selected. In Section 4 we classify each of the relevant studies according to novel types of miscategorisation bias exhibited. In Section 5 we simulate the vaccine efficacy results that would be observed during peak rollout of both a placebo and negative efficacy vaccine under the various miscategorisation biases. Section 6 offers our conclusions.

## 2. Background

Several studies have investigated bias in Covid-19 vaccine studies, including: (i) *outcome reporting bias* affecting interpretation of vaccine efficacy where studies report relative risk reduction (RRR) rather than actual risk reduction (ARR) (Brown, 2021); *confounding bias* in test-negative studies where other acute respiratory infections (ARI) are assumed to occur or be independent to Covid-19 (Doll et al, 2022), where authors promote the use of recently vaccinated individuals as a negative control (Hitchings et al, 2022), due to imperfect sensitivity and/or specificity of the test used to diagnose the disease (Eusebi et al, 2023; Williams et al, 2022); *state bias* wherein limited uptake, or vaccine hesitancy, is said to occur because the general public prefer domestically produced vaccines over foreign-made (Kobayashi et al, 2021) and alternatively, *confirmation bias* that causes people to disregard public information and results in the same hesitancy (Malthouse, 2023); *self-selection bias* where participants who have been vaccinated are more likely to also willingly present for swab collection and testing (Glasziou et al, 2022); and *collider stratification bias* where rather than the usual approach of reporting the *relative risk of the disease*, Covid-19, test-negative studies use the recently created alternative approach of reporting the *relative risk of infection* given a second variable, vaccination (Ortiz-Brizuela et al, 2023). The studies discussed here are approximately evenly divided between those that report biases that have exaggerated factors of vaccine safety and efficacy, and those reporting biases that have negatively impacted assessment of these factors and resulting public perception.

A consistent bias in studies of Covid-19 vaccine effectiveness arises from the assumption that it is necessary to allow an incubation period (typically up to 21 days) for an immune response to take effect. Under this assumption subjects are not categorised as vaccinated until this period has lapsed. This is justified, for example in (Polack et al, 2020), with data that indicate that a divergence in Covid-19 cases between the vaccinated and unvaccinated only occurs after at least 12 days. However, in (Lauer et al, 2020) the authors admit that “Our current understanding of the incubation period for COVID-19 is limited”. From observational studies, such as (Pilishvili et al, 2021), it is known that a disproportionately larger number of Covid-19 cases are detected, in the vaccinated cohort compared to the unvaccinated cohort, within the first 10-14 days after vaccination. They reported that for the period of 0-9 days after receipt of the first dose, vaccine effectiveness was 12.8% and vaccine effectiveness at 10-13 days after receipt of the first dose was 36.8%.

Clearly, this is not an indication of an ‘effective vaccine’. Indeed, we might consider a hypothetical scenario where this incubation period assumption operates and where every vaccinated person is infected within the first 21 days yet is not categorised as vaccinated and is instead categorised as unvaccinated. Logically, if at least one genuinely unvaccinated person is infected within the period of the study, we would then conclude the vaccine, despite offering zero protection against infection, is 100% effective.

We focus explicitly on miscategorisation biases, which in the Covid-19 studies cited have inevitably exaggerated vaccine efficacy. We identify five types of such bias (defined in detail in Section 4), namely: (i) *Basic miscategorisation* (the type most closely associated with the miscategorisation bias); (ii) *Unverified*; (iii) *Uncontrolled*; (iv) *Excluded*; and (v) *Undefined*. Previous work (Ioannidis, 2021; Fung, Jones & Doshi, 2023, Lataster 2024) has largely focused only on miscategorisation, so our review is novel as well as more extensive than previous work. Ioannidis (2021) considers miscategorisation in terms of vaccination self-reporting by participants, the need for investigators to provide definitions for what it means to be vaccinated and whether categorisation as vaccinated occurs immediately after vaccination or after some period, and they discuss the possibility for these definitions to themselves cause miscategorisation of vaccination status. Fung et al (2023) examine this issue in terms of a case-counting window bias, in which investigators do not begin counting cases in the *fully vaccinated* until the arbitrary period after vaccination had passed. They also found that investigators could apply this period to both the vaccine and placebo arms of their study, or to the vaccine group alone.

## 3 Method

A search was conducted of PubMed and Scopus seeking literature presenting either a retrospective health records or prospective clinical trial of one or more Covid-19 vaccines with efficacy or safety as an endpoint. The search term used was:

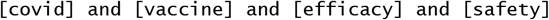

For inclusion, a paper had to provide a primary report of either a prospective or retrospective Covid-19 vaccination trial comparing infection rates, with or without adverse events, between vaccinated and unvaccinated individuals. Works were excluded if they were: (a) not reporting a vaccine trial or infection rate comparison pre and post vaccination; (b) not reporting a control or unvaccinated group; (c) a review of other works; (d) a response to or letter to the editor about a vaccine trial; (e) a protocol for a proposed vaccine trial or comparison; or (f) any other form of non-peer reviewed discourse. We included both safety and efficacy as keywords because often studies made implicit claims about safety based on an evaluation of efficacy without any formal justification.

The initial search returned 2,209 results. Four additional works were identified through citation mining. 476 Duplicates were removed. Another 1,697 were removed: 1,561 that while discussing or mentioning vaccines for Covid-19 did not present a study of vaccine efficacy or safety, and 136 single-page works that were a mix of protocol disclosures and abstracts of results. Of the 40 remaining, three additional papers were excluded because they did not use miscategorisation and are out of scope for this study (Baden et al, 2020; Khairullin et al, 2020; Polack et al, 2020). This left 37 that provide sufficient detail of the inclusion and exclusion criteria for inclusion in this study. Each paper was evaluated for a range of aspects that included the manufacturer and type of vaccine, the control cohort comparator (placebo or unvaccinated), the primary outcomes (prevention of infection, hospitalisation, ICU admission or death), the author’s potential conflicts of interest (declared and undeclared) and whether they included one or more types of miscategorisation bias. This work reports on the last of these factors.

The PRISMA (Preferred Reporting Items for Systematic Reviews and Meta-Analyses) diagram for this literature search is shown in Figure 1.

**Figure 1:**
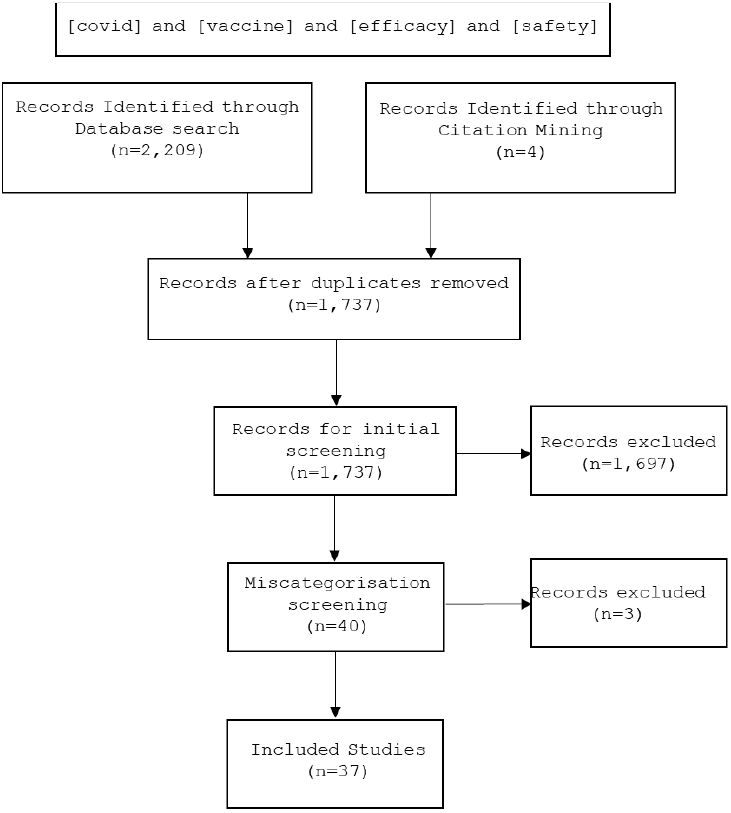
PRISMA Diagram.

## 4. Types of miscategorisation bias

Previous papers identifying biases in the Covid-19 vaccine studies did not provide a methodology for the approach they used to review and identify either the biases themselves, or the works that contained those biases (Fung, Jones & Doshi, 2023; Ioannidis, 2022). There are a range of known issues that can affect clinical trial studies that include the degree to which participants and clinicians are blinded, whether confounding variables have been identified and controlled for, and whether participants are correctly identified into appropriate groups. Works, such as Rothman et al (2008) discuss a range of these issues that can arise in clinical studies and lead to bias, which we used to develop a list of issues for identification in the reviewed Covid-19 vaccine studies. We have grouped these together under the following five types of the miscategorisation bias:

*(a) Basic miscategorisation:* During the arbitrarily defined period the vaccinated are categorised as unvaccinated, twice vaccinated categorised as single vaccinated, or boosted categorised as twice vaccinated (e.g.: Buchan et al, 2022; Stock et al, 2022).

*(b) Unverified:* Participants whose vaccination status is unknown or unverified are categorised as unvaccinated (e.g.: Rosenberg et al, 2021; Lyngse et al, 2022b).

*(c) Uncontrolled:* Participants are allowed to self-administer or self-report their vaccination or infection status, became unblinded or sought vaccination outside the study (e.g.: Angel et al, 2021).

*(d) Excluded:* Participants who are vaccinated but who become infected or died during the arbitrarily defined period are categorised as neither unvaccinated or vaccinated but are instead simply removed from analysis (e.g.: Tabarsi et al, 2023; Heath et al, 2023). Note that there are two forms of exclusion applied, symmetric (S) where participants are excluded on all arms of the study, including the control arm, and asymmetric (A) where they are excluded only in the treatment arm.

*(e) Undefined:* The authors of the study fail to provide definitions for either or both vaccinated and unvaccinated cohorts (e.g.: Bermingham et al, 2023b; Nordstrom et al, 2022).

Table 1 lists the incidence and frequency of use for each type of miscategorisation bias in Covid-19 vaccine effectiveness research studies. Use of the arbitrary miscategorisation type was ubiquitous, identified in 100% of the reviewed studies. Further, nearly one-third also used one or more of the other types of bias.

**Table 1.** Research studies containing miscategorisation bias (see appendix for full citation list)

| **Citation** | **(a)** | **(b)** | **(c)** | **(d)** | **(e)** | **Defined Period** |
| --- | --- | --- | --- | --- | --- | --- |
| Dagan et al (2021) | X |  |  |  |  | 14 days |
| Haas et al (2021) | X |  |  |  |  | 7 days |
| Rosenberg et al (2021) | X | X |  |  |  | 14 days |
| Thomas et al (2021) | X |  |  |  |  | 7 days |
| Angel et al (2021) | X |  | X |  |  | 7 days |
| NSW Health (2021) | X | X |  |  |  | 14 days |
| Ali et al (2021) | X |  |  |  |  | 14 days |
| Pilishvili et al (2021) | X |  | X |  |  | 14 days / 7 days |
| Andrews et al (2022) | X |  |  |  |  | 28 days |
| Buam et al (2022) | X |  |  |  |  | 21 days / 14 days |
| Buchan et al (2022) | X |  |  |  |  | 7 days |
| Carazo et al (2022) | X |  |  |  |  | 14 days |
| Chung et al (2022) | X |  |  |  |  | 7 days |
| Palinkas et al (2022) | X |  |  |  |  | 7 days |
| Ferdinands et al (2022) | X | X |  | X (A) |  | 14 days |
| Lyngse et al (2022) | X |  |  |  |  | 7-15 days |
| Lyngse et al (2022b) | X | X |  |  |  | 7-15 days |
| Nordstrom et al (2022) | X |  |  | X (A) | X | 14 days |
| Petras et al (2022) | X |  |  |  |  | 14 days |
| Robles-Fontan et al (2022) | X |  |  |  |  | 14 days |
| Arbel et al (2022) | X |  |  |  |  | 7 days |
| Paternina et al (2022) | X |  |  |  |  | 14 days |
| Stock et al (2022) | X |  |  |  |  | 21 days / 14 days |
| Birmingham et al (2023) | X |  |  |  |  | 21 days |
| Yau et al (2023) | X |  |  |  |  | Until fully vaccinated |
| Mitchell et al (2023) | X |  |  |  |  | 14 days |
| Tan et al (2023) | X |  |  |  |  | 7 days |
| Al Kaabi et al (2023) [RCT] |  |  |  | X (A) |  | 14 days |
| Tabarsi et al (2023) [RCT] |  |  |  | X (S) |  | 14 days |
| Heath et al (2023) [RCT] |  |  |  | X (A) |  | 7 days |
| Nadeem et al (2023) | X |  |  |  |  | 14 days |
| Anez et al (2023) [RCT] |  |  |  | X (S) |  | 7 days |
| Munoz et al (2023) [RCT] |  |  |  | X (S) |  | 14 days |
| Birmingham et al (2023b) | X |  |  |  | X | 21 days |
| Liu et al (2023) | X |  |  |  |  | 7 days |
| Kitano et al (2023) | X |  |  |  |  | 7 days / 14 days |
| Wu et al (2023) |  |  |  | X(A) |  | 28 days |
|  | **31** | **4** | **2** | **8** | **2** |  |

## 5. Simulation of vaccine effectiveness

We used a deterministic temporal simulation to illustrate the effects of the miscategorisation bias on vaccine effectiveness and the reported infection rates for different cohorts, vaccinated and unvaccinated. We simulated a hypothetical vaccination campaign starting at week 1 and completing on week 6 with 85% of the observed population vaccinated by that time.

Here we examine several scenarios showing the effect of a one-week, two-week and three-week biases for miscategorisation (a) and asymmetric exclusion (d) and the effects of repeated vaccination, by boosting, on vaccine efficacy and infection reported rates. Two scenarios present a placebo (zero- efficacy) vaccine, which does not affect infection rates, and compare this with a negative-efficacy vaccine, whereby those vaccinated suffer slightly elevated infection rates compared to the unvaccinated. We also include a protective, positive-efficacy, vaccine scenario for comparison.

Note that observational studies and randomised control trials might suffer from many sources of additional confounding biases so this model is a simplification and should not be taken as representative of population level data.

The scenarios simulated cover an eleven-week period with an assumed constant weekly infection rate of 1% in the placebo scenario, and a slightly elevated infection rate, 1.25%, for the vaccinated cohort in the negative-efficacy scenario. This is used in both the miscategorisation, (a), and asymmetrically excluded, (d), simulations. To simulate the effects of boosters we assume a population that is repeatedly vaccinated every twelve weeks, with those who are vaccinated miscategorised (a) within one week of each vaccination. In the protective (positive efficacy) scenarios, E and F, we assume the weekly infection rate for those vaccinated within 1, 2 or 3 weeks since vaccination is double that for the period after, at 0.5% when supposed immunity is reached and 1% beforehand (i.e. the same as the unvaccinated). Note that we assume that previously infected members of the simulated population can become reinfected and hence do not become immune. Also, for low infection rates, as we model here, the difference between assuming immunity and not, is negligible.

In Scenario G, we examine the effects of boosting with miscategorisation, (a), to determine whether repeated application of the vaccine at twelve-week intervals restores vaccine efficacy to high levels after each booster and whether it elevates the reported infection rate in the unvaccinated cohort between each booster campaign.

In scenario H we look at symmetric exclusion of cases across both trial arms. Here we assume the same infection rate for the unvaccinated and vaccinated, 1%, except that during the duration of the exclusion period the vaccinated suffer a temporary increase in infection of 2%, reflecting a detrimental effect of the vaccine on the immune system. Given that symmetric exclusion can only occur in randomised control trials, we have assumed identical populations in both trial arms with vaccinated and unvaccinated participants matched together.

The method for calculating vaccine effectiveness and all scenario results are provided in supplementary materials. Note that two methods of calculating the vaccine effectiveness denominators are possible, one where the population denominators are adjusted to remove those cases excluded from, or transferred between, categories and one where they are not. The latter method (denominators unadjusted) is the convention, and our results are calculated using that method. Note that when the infection rates are low, and the population is large one method approximates the other. It is also important to note that, because the focus in observational studies is on cases, typically only these people in the population are classified as unvaccinated if they happen within the delay period. The simulations reflect this practice.

The results of the eight scenarios are presented in Figure 2.

**Figure 2.**
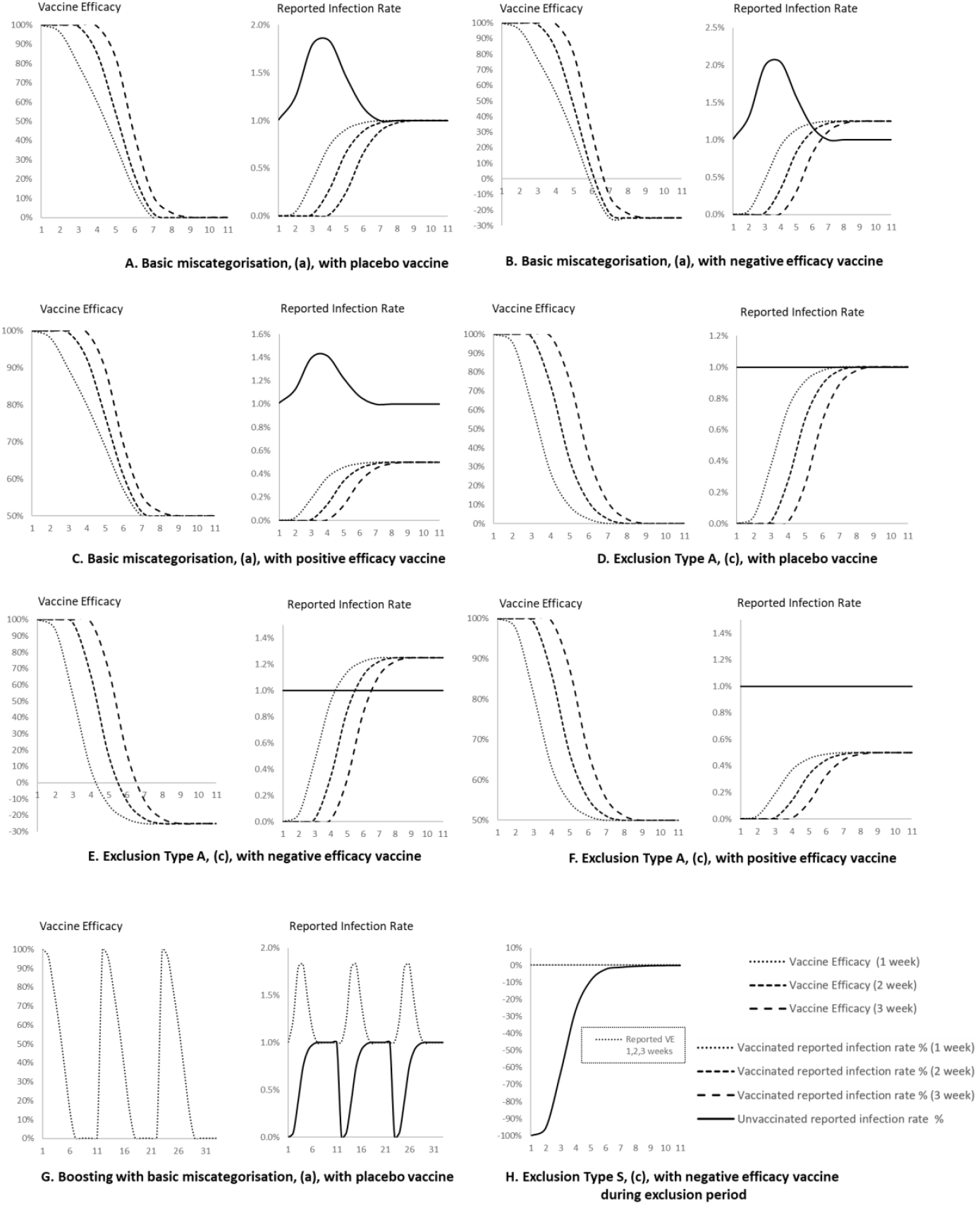
Eight scenarios A-H. A: Basic miscategorisation, (a) with placebo vaccine; B: Basic miscategorisation, (a), with negative efficacy vaccine; C: Basic miscategorisation, (a), with positive efficacy vaccine; D: Exclusion (type A), (c) with placebo vaccine; E: Exclusion (type A), (c), with negative efficacy vaccine; F: Exclusion (type A), (c), with positive efficacy vaccine; G: Boosting with basic miscategorisation, (a), with placebo vaccine; H: Exclusion (type S) (c), with negative efficacy vaccine during exclusion period.

In practice, most studies do not report vaccine efficacy in the initial week(s) (when no cases are categorised as vaccinated) as this would show up as 100% efficacy. However, note that in all scenarios in the first weeks where efficacy would be reported the starting point for reported efficacy is over 90%. Scenarios D, E and F are simply the same as scenarios A, B and C, except for the fact that they are for the asymmetrically excluded type, (c), of miscategorisation bias. Note that here the reported infection rate for the unvaccinated remains unbiased whilst that for the vaccinated rises to match the true rate for the placebo and negative efficacy scenarios.

In scenario A, miscategorisation, (a), with a placebo, high vaccine effectiveness falls towards zero after one, two or three-week periods, accompanied by an increase in the reported infection rate for the unvaccinated cohort from the start of the vaccination campaign. After seven weeks the reported infection rates for the vaccinated and unvaccinated cohorts converge on the true infection rate. In scenario B, miscategorisation, (a), with a negative effectiveness vaccine, the reported vaccine effectiveness is negative from week six onwards, and again the reported infection rate for the unvaccinated is overestimated from the start of the vaccination campaign. However, by the end of the campaign the reported infection rates for the vaccinated would be greater than that for the unvaccinated. In scenario C, miscategorisation, (a), with a positive efficacy vaccine, vaccine effectiveness falls to 50% rather than 0% or below 0% as it does for scenarios A and B and will never decrease below this.

In scenario A the *reported* infection rate for the unvaccinated increases because the number of people infected post vaccination, at any point in the previous 1, 2 or 3 weeks is added to the number of unvaccinated people infected who are unvaccinated that week. Thus, despite the actual infection rates being the same for both groups, we add more than a single weeks’ worth of infected cases from the vaccinated group to a single weeks’ worth of infected cases in the unvaccinated group. This gives rise to an artificial rise in the *reported* infection rate compared to the *true* infection rate (1% here). This same effect occurs in scenarios B and C.

Likewise, in scenario A, during the initial 1,2- or 3-week period the number of vaccinated infected are not reported as infected but are instead reported as having occurred in the unvaccinated group, hence the reported infection rate for those vaccinated starts at zero and then climbs until it converges with the true infection rate. In scenario B, where we have a negative efficacy vaccine, the reported infection rate for the vaccinated is under-reported until it converges with the actual vaccinated infection rate, which is higher than the infection rate for the unvaccinated. In scenario C, where we have positive efficacy, the infection rate for the vaccinated is again under-reported until it converges to the infection rate conferred by immunisation.

Scenarios D, E and F cover asymmetrical exclusion, (c) with placebo, negative efficacy and positive efficacy vaccines. These scenarios reveal that the major difference, compared with the miscategorisation scenarios, is that the reported infection rate for the unvaccinated is never over-reported, whilst the reported infection rate for the vaccinated is over-reported, and thus exaggerates the actual protective benefit of the vaccine and vaccine efficacy.

In Scenario G, boosting with miscategorisation, (a), we can see that repeated application of the vaccine at twelve-week intervals restores vaccine efficacy to high levels after each booster and, assuming a constant infection rate, elevates the reported infection rate in the unvaccinated cohort between each booster campaign, giving rise to bias and gross overestimation.

Scenario H covers symmetric exclusion, (c), with a negative efficacy vaccine whose detrimental effect on the immune system only occurs during the period of exclusion. We can see that without the practice of symmetric exclusion of both cases from the vaccinated and unvaccinated cohorts the actual vaccine efficacy would start at −100% and rise towards zero, whereas the reported vaccine efficacy would be zero throughout the reported period, thus completely masking the negative effects of the vaccine. To achieve positive efficacy other adjustments to how cases were handled would be required, such as biases in symptom screening or testing rates.

Our simulation model has demonstrated that the effects of this miscategorisation bias are to artificially boost vaccine efficacy in all cases, and with the application of repeated ‘booster’ vaccinations, the efficacy of repeated Covid-19 vaccines could be maintained at these artificial levels in perpetuity should boosting be continued indefinitely. Furthermore, in tandem with this the infection rate is likewise artificially elevated for the unvaccinated cohort compared to the vaccinated cohort, further compounding false claims that a Covid-19 vaccine reduces infection rates. Note that other metrics of vaccine effectiveness, such as mortality or morbidity improvements, are capable of being mis-reported in a similar way because of the same bias.

## 6. Conclusions

Our review reveals that a serious form of bias, miscategorisation, is pervasive throughout the many research studies that aim to measure Covid-19 vaccine efficacy. The effect of this bias is to artificially inflate vaccine efficacy and present the misleading impression that these vaccines are effective and that the non-vaccinated suffer from higher Covid-19 infection rates compared to the vaccinated.

We presented a simulation model to demonstrate the effects of this bias and show it artificially boosts vaccine efficacy in all cases, and with the application of repeated ‘booster’ vaccinations, the efficacy of repeated Covid-19 vaccines could be maintained at artificial levels in perpetuity should boosting be continued indefinitely. This effect occurs with a both a zero-efficacy (placebo) vaccine and a negative- efficacy vaccine that increases, rather than reduces, infection rates in those vaccinated.

This miscategorisation is guaranteed to lead to initially very high efficacy claims (usually above 90%) during peak vaccine rollout even if the vaccine were a placebo or worse. Efficacy then falls toward zero a few weeks later. This pattern of high initial efficacy, tapering off after 3 months is also consistently observed in real-world studies, and is often used as justification for additional, booster vaccinations to maintain efficacy. The corresponding Covid-19 infection rate is also likewise artificially elevated in the unvaccinated cohort compared to the vaccinated cohort. These issues apply to other measures of vaccination effectiveness related to mortality and morbidity.

Thus, we conclude that any claims of Covid-19 vaccine efficacy based on these studies are likely to be a statistical illusion or are exaggerated.

## Data Availability

All relevant data is contained within the manuscript

## Supplementary materials

**VE (vaccine efficacy) calculation:**

*VE*_*t*_ is the efficacy of a vaccine, at time *t*, expressed as a percentage.

Miscategorisation, not including denominator change:

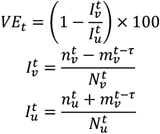

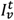 is the reported infection rate for the cumulative ever vaccinated population, *v*, at time *t*.

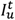 is the reported infection rate for the cumulative unvaccinated population, *u*, at time *t*.

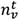 is the cumulative number of infected or reinfected vaccinated population at time *t*.

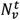 is the cumulative number of vaccinated population up to time *t*.

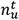 is the cumulative number of infected or reinfected unvaccinated population at time *t*.

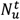 is the cumulative number of unvaccinated population up to time *t*.

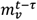 is the cumulative number of ‘newly’ vaccinated population infected within *t* less *τ* weeks since vaccination who are categorised as unvaccinated at time *t* where *τ* = 1,2,3 and *t* − *τ* > 0.

If denominator change is accommodated, then:

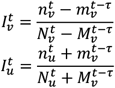

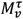 is the total population of ‘newly’ vaccinated that are infected within *t* less than *τ* weeks of vaccination who are categorised as unvaccinated up to time *t* where *τ* = 1,2,3 and *t* − *τ* > 0 and where:

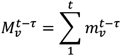

Exclusion, not including denominator change:

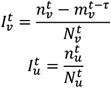

Exclusion, with denominator change:

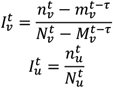

## Simulation Results

### Scenario A – miscategorisation + placebo vaccine

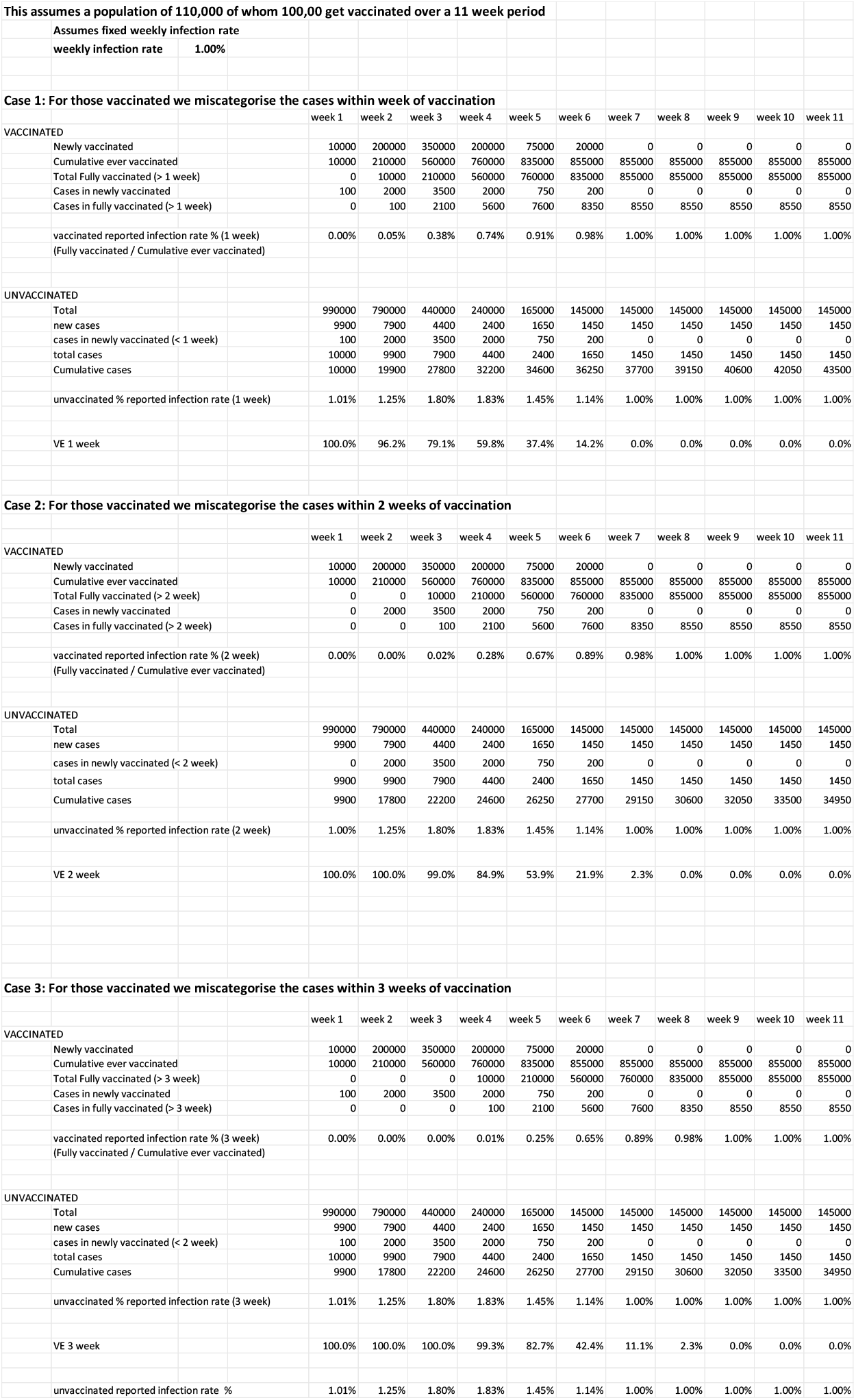

### Scenario B miscategorisation + negative efficacy vaccine

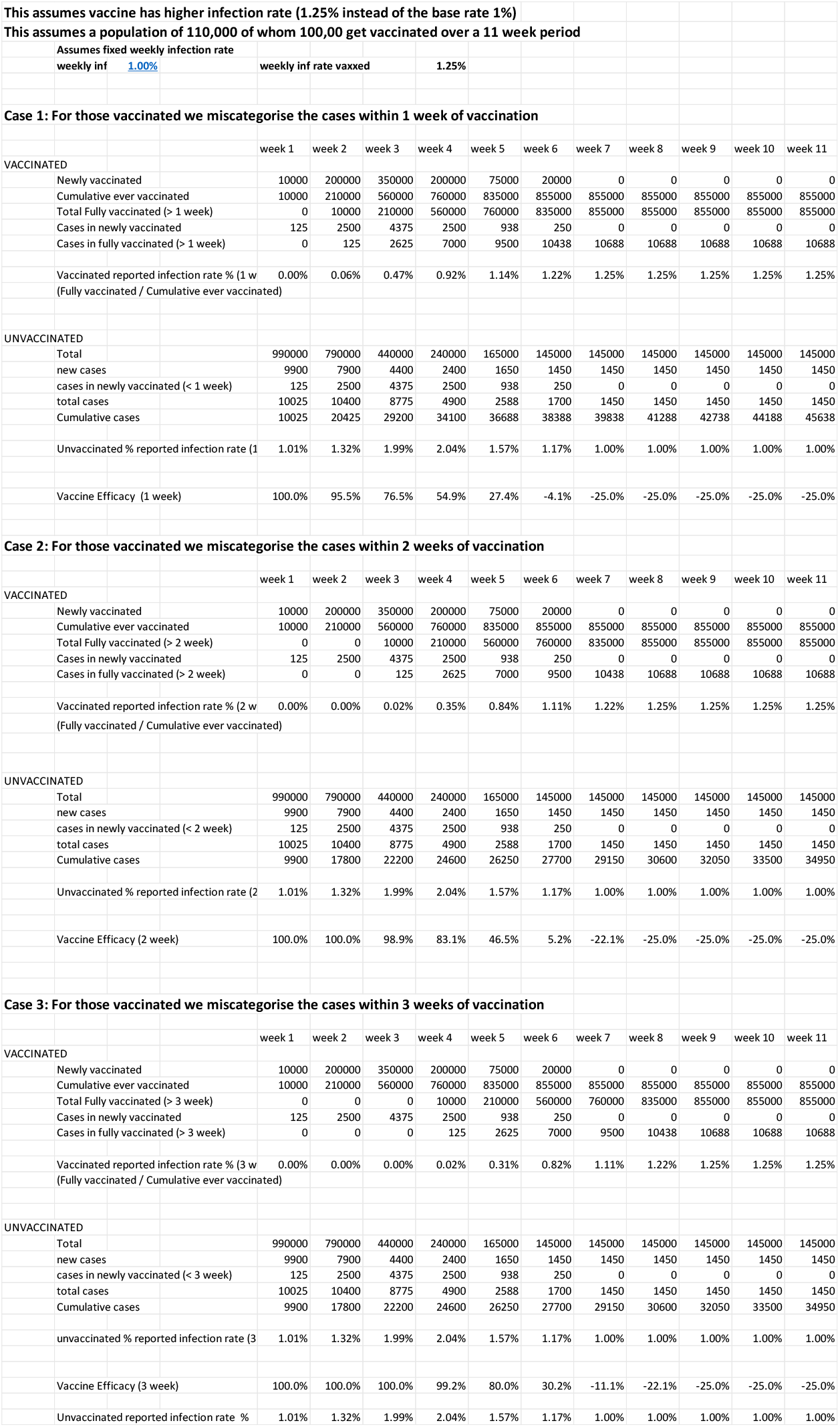

### Scenario C miscategorisation + positive efficacy vaccine

| this assumes a population of 110,000 of whom 100,00 get vaccinated over a 11 week period |  |  |  |  |  |  |  |  |  |  |  |  |  |  |
| --- | --- | --- | --- | --- | --- | --- | --- | --- | --- | --- | --- | --- | --- | --- |
| Assumes fixed weekly infection rate |  | weekly inf |  | 1.00% |  | weekly inf rate vaxxed |  | 0.50% |  |  |  |  |  |  |
| Case 1: For those vaccinated we miscategorise the cases within 1 week of vaccination |  |  |  |  |  |  |  |  |  |  |  |  |  |  |
|  |  |  |  | week 1 | week 2 | week 3 | week 4 | week 5 | week 6 | week 7 | week 8 | week 9 | week 10 | week 11 |
| VACCINATED |  |  |  |  |  |  |  |  |  |  |  |  |  |  |
|  | Newly vaccinated |  |  | 10000 | 200000 | 350000 | 200000 | 75000 | 20000 | 0 | 0 | 0 | 0 | 0 |
|  | Cumulative ever vaccinated |  |  | 10000 | 210000 | 560000 | 760000 | 835000 | 855000 | 855000 | 855000 | 855000 | 855000 | 855000 |
|  | Total Fully vaccinated (> 1 week) |  |  | 0 | 10000 | 210000 | 560000 | 760000 | 835000 | 855000 | 855000 | 855000 | 855000 | 855000 |
|  | Cases in newly vaccinated |  |  | 100 | 2000 | 3500 | 2000 | 750 | 200 | 0 | 0 | 0 | 0 | 0 |
|  | Cases in fully vaccinated (> 1 week) |  |  | 0 | 50 | 1050 | 2800 | 3800 | 4175 | 4275 | 4275 | 4275 | 4275 | 4275 |
|  | Vaccinated reported infection rate % (1 w<br>(Fully vaccinated / Cumulative ever vaccinated) |  |  | 0.00% | 0.02% | 0.19% | 0.37% | 0.46% | 0.49% | 0.50% | 0.50% | 0.50% | 0.50% | 0.50% |
| UNVACCINATED |  |  |  |  |  |  |  |  |  |  |  |  |  |  |
|  | Total |  |  | 990000 | 790000 | 440000 | 240000 | 165000 | 145000 | 145000 | 145000 | 145000 | 145000 | 145000 |
|  | new cases |  |  | 9900 | 7900 | 4400 | 2400 | 1650 | 1450 | 1450 | 1450 | 1450 | 1450 | 1450 |
|  | cases in newly vaccinated (< 1 week) |  |  | 100 | 2000 | 3500 | 2000 | 750 | 200 | 0 | 0 | 0 | 0 | 0 |
|  | total cases |  |  | 10000 | 9900 | 7900 | 4400 | 2400 | 1650 | 1450 | 1450 | 1450 | 1450 | 1450 |
|  | Cumulative cases |  |  | 10000 | 19900 | 27800 | 32200 | 34600 | 36250 | 37700 | 39150 | 40600 | 42050 | 43500 |
|  | Unvaccinated % reported infection rate (1 |  |  | 1.01% | 1.25% | 1.80% | 1.83% | 1.45% | 1.14% | 1.00% | 1.00% | 1.00% | 1.00% | 1.00% |
|  | Vaccine Efficacy (1 week) |  |  | 100.0% | 98.1% | 89.6% | 79.9% | 68.7% | 57.1% | 50.0% | 50.0% | 50.0% | 50.0% | 50.0% |
| Case 2: For those vaccinated we miscategorise the cases within 2 weeks of vaccination |  |  |  |  |  |  |  |  |  |  |  |  |  |  |
|  |  |  |  | week 1 | week 2 | week 3 | week 4 | week 5 | week 6 | week 7 | week 8 | week 9 | week 10 | week 11 |
| VACCINATED |  |  |  |  |  |  |  |  |  |  |  |  |  |  |
|  | Newly vaccinated |  |  | 10000 | 200000 | 350000 | 200000 | 75000 | 20000 | 0 | 0 | 0 | 0 | 0 |
|  | Cumulative ever vaccinated |  |  | 10000 | 210000 | 560000 | 760000 | 835000 | 855000 | 855000 | 855000 | 855000 | 855000 | 855000 |
|  | Total Fully vaccinated (> 2 week) |  |  | 0 | 0 | 10000 | 210000 | 560000 | 760000 | 835000 | 855000 | 855000 | 855000 | 855000 |
|  | Cases in newly vaccinated |  |  | 100 | 2000 | 3500 | 2000 | 750 | 200 | 0 | 0 | 0 | 0 | 0 |
|  | Cases in fully vaccinated (> 2 week) |  |  | 0 | 0 | 50 | 1050 | 2800 | 3800 | 4175 | 4275 | 4275 | 4275 | 4275 |
|  | Vaccinated reported infection rate % (2 w<br>(Fully vaccinated / Cumulative ever vaccinated) |  |  | 0.00% | 0.00% | 0.01% | 0.14% | 0.34% | 0.44% | 0.49% | 0.50% | 0.50% | 0.50% | 0.50% |
| UNVACCINATED |  |  |  |  |  |  |  |  |  |  |  |  |  |  |
|  | Total |  |  | 990000 | 790000 | 440000 | 240000 | 165000 | 145000 | 145000 | 145000 | 145000 | 145000 | 145000 |
|  | new cases |  |  | 9900 | 7900 | 4400 | 2400 | 1650 | 1450 | 1450 | 1450 | 1450 | 1450 | 1450 |
|  | cases in newly vaccinated (< 2 week) |  |  | 100 | 2000 | 3500 | 2000 | 750 | 200 | 0 | 0 | 0 | 0 | 0 |
|  | total cases |  |  | 10000 | 9900 | 7900 | 4400 | 2400 | 1650 | 1450 | 1450 | 1450 | 1450 | 1450 |
|  | Cumulative cases |  |  | 9900 | 17800 | 22200 | 24600 | 26250 | 27700 | 29150 | 30600 | 32050 | 33500 | 34950 |
|  | Unvaccinated % reported infection rate (2 |  |  | 1.01% | 1.25% | 1.80% | 1.83% | 1.45% | 1.14% | 1.00% | 1.00% | 1.00% | 1.00% | 1.00% |
|  | Vaccine Efficacy (2 week) |  |  | 100.0% | 100.0% | 99.5% | 92.5% | 76.9% | 60.9% | 51.2% | 50.0% | 50.0% | 50.0% | 50.0% |
| Case 3: For those vaccinated we miscategorise the cases within 3 weeks of vaccination |  |  |  |  |  |  |  |  |  |  |  |  |  |  |
|  |  |  |  | week 1 | week 2 | week 3 | week 4 | week 5 | week 6 | week 7 | week 8 | week 9 | week 10 | week 11 |
| VACCINATED |  |  |  |  |  |  |  |  |  |  |  |  |  |  |
|  | Newly vaccinated |  |  | 10000 | 200000 | 350000 | 200000 | 75000 | 20000 | 0 | 0 | 0 | 0 | 0 |
|  | Cumulative ever vaccinated |  |  | 10000 | 210000 | 560000 | 760000 | 835000 | 855000 | 855000 | 855000 | 855000 | 855000 | 855000 |
|  | Total Fully vaccinated (> 3 week) |  |  | 0 | 0 | 0 | 10000 | 210000 | 560000 | 760000 | 835000 | 855000 | 855000 | 855000 |
|  | Cases in newly vaccinated |  |  | 100 | 1000 | 1750 | 1000 | 375 | 100 | 0 | 0 | 0 | 0 | 0 |
|  | Cases in fully vaccinated (> 3 week) |  |  | 0 | 0 | 0 | 50 | 1050 | 2800 | 3800 | 4175 | 4275 | 4275 | 4275 |
|  | Vaccinated reported infection rate % (3 w<br>(Fully vaccinated / Cumulative ever vaccinated) |  |  | 0.00% | 0.00% | 0.00% | 0.01% | 0.13% | 0.33% | 0.44% | 0.49% | 0.50% | 0.50% | 0.50% |
| UNVACCINATED |  |  |  |  |  |  |  |  |  |  |  |  |  |  |
|  | Total |  |  | 990000 | 790000 | 440000 | 240000 | 165000 | 145000 | 145000 | 145000 | 145000 | 145000 | 145000 |
|  | new cases |  |  | 9900 | 7900 | 4400 | 2400 | 1650 | 1450 | 1450 | 1450 | 1450 | 1450 | 1450 |
|  | cases in newly vaccinated (< 3 week) |  |  | 100 | 1000 | 1750 | 1000 | 375 | 100 | 0 | 0 | 0 | 0 | 0 |
|  | total cases |  |  | 10000 | 8900 | 6150 | 3400 | 2025 | 1550 | 1450 | 1450 | 1450 | 1450 | 1450 |
|  | Cumulative cases |  |  | 9900 | 17800 | 22200 | 24600 | 26250 | 27700 | 29150 | 30600 | 32050 | 33500 | 34950 |
|  | unvaccinated % reported infection rate (3 |  |  | 1.01% | 1.13% | 1.40% | 1.42% | 1.23% | 1.07% | 1.00% | 1.00% | 1.00% | 1.00% | 1.00% |
|  | Vaccine Efficacy (3 week) |  |  | 100.0% | 100.0% | 100.0% | 99.5% | 89.8% | 69.4% | 55.6% | 51.2% | 50.0% | 50.0% | 50.0% |
|  | Unvaccinated reported infection rate % |  |  | 1.01% | 1.13% | 1.40% | 1.42% | 1.23% | 1.07% | 1.00% | 1.00% | 1.00% | 1.00% | 1.00% |

### Scenario D – asymmetric exclusion + placebo vaccine

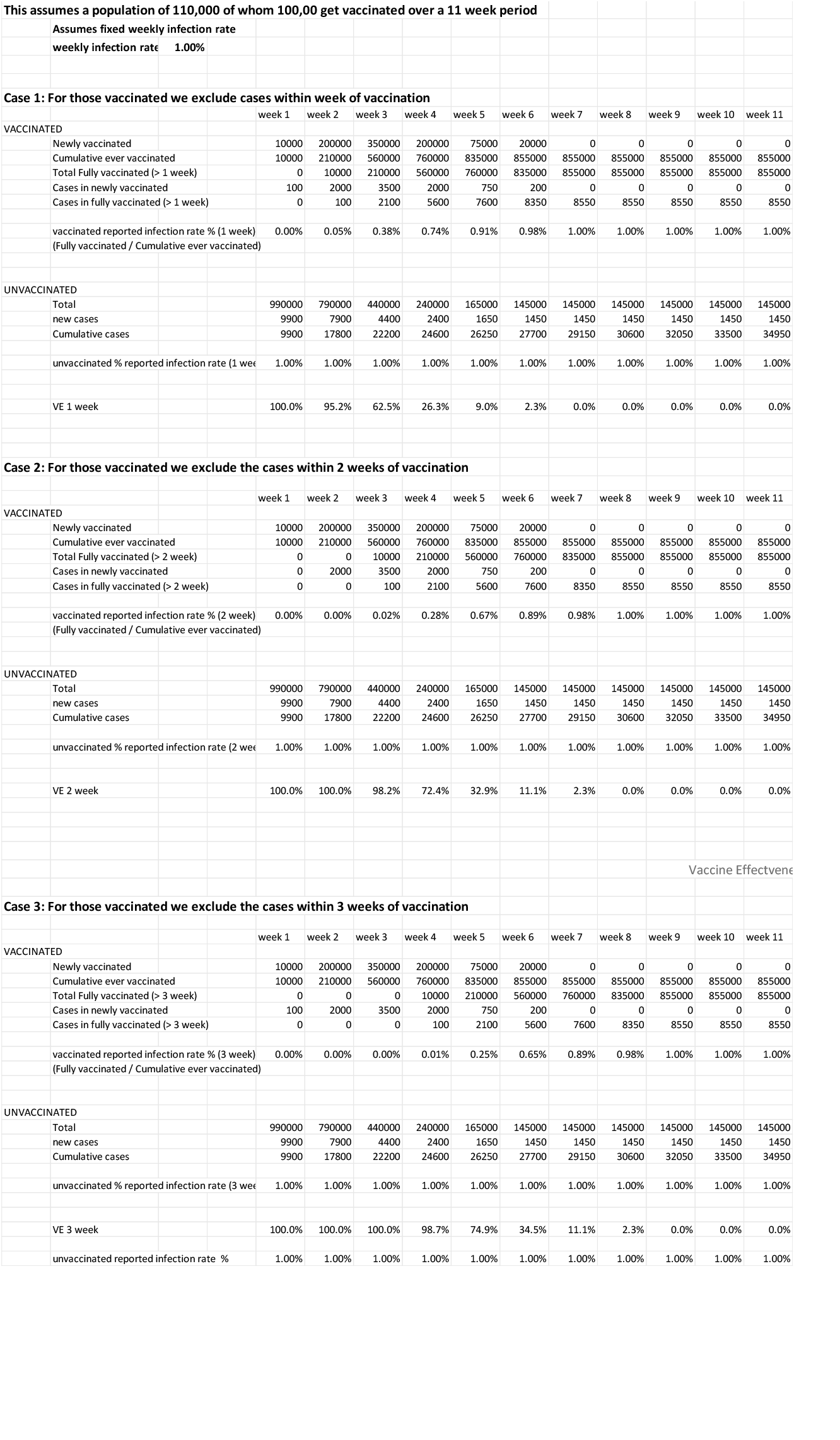

### Scenario E – asymmetric exclusion + negative efficacy vaccine

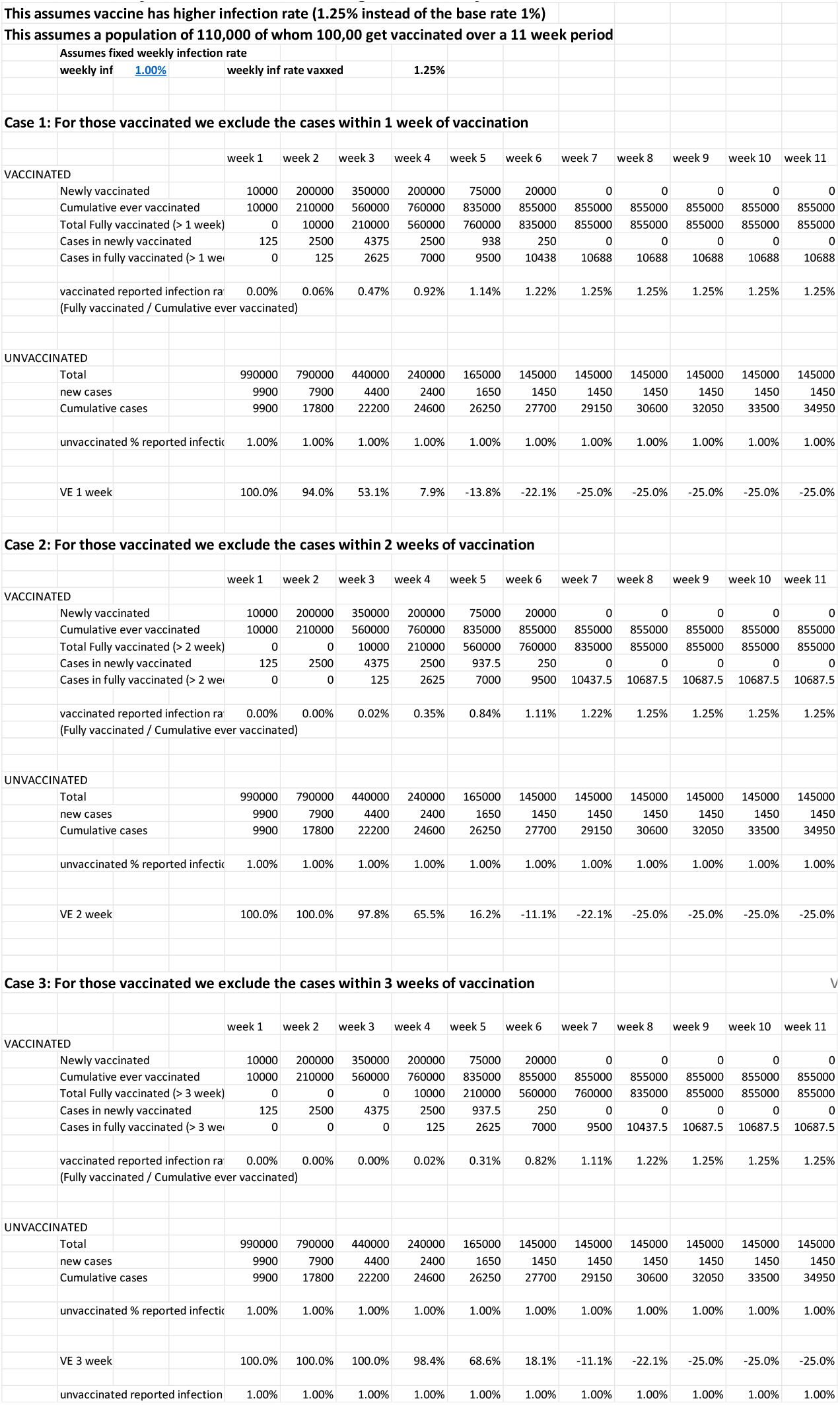

### Scenario F – asymmetric exclusion + positive efficacy vaccine

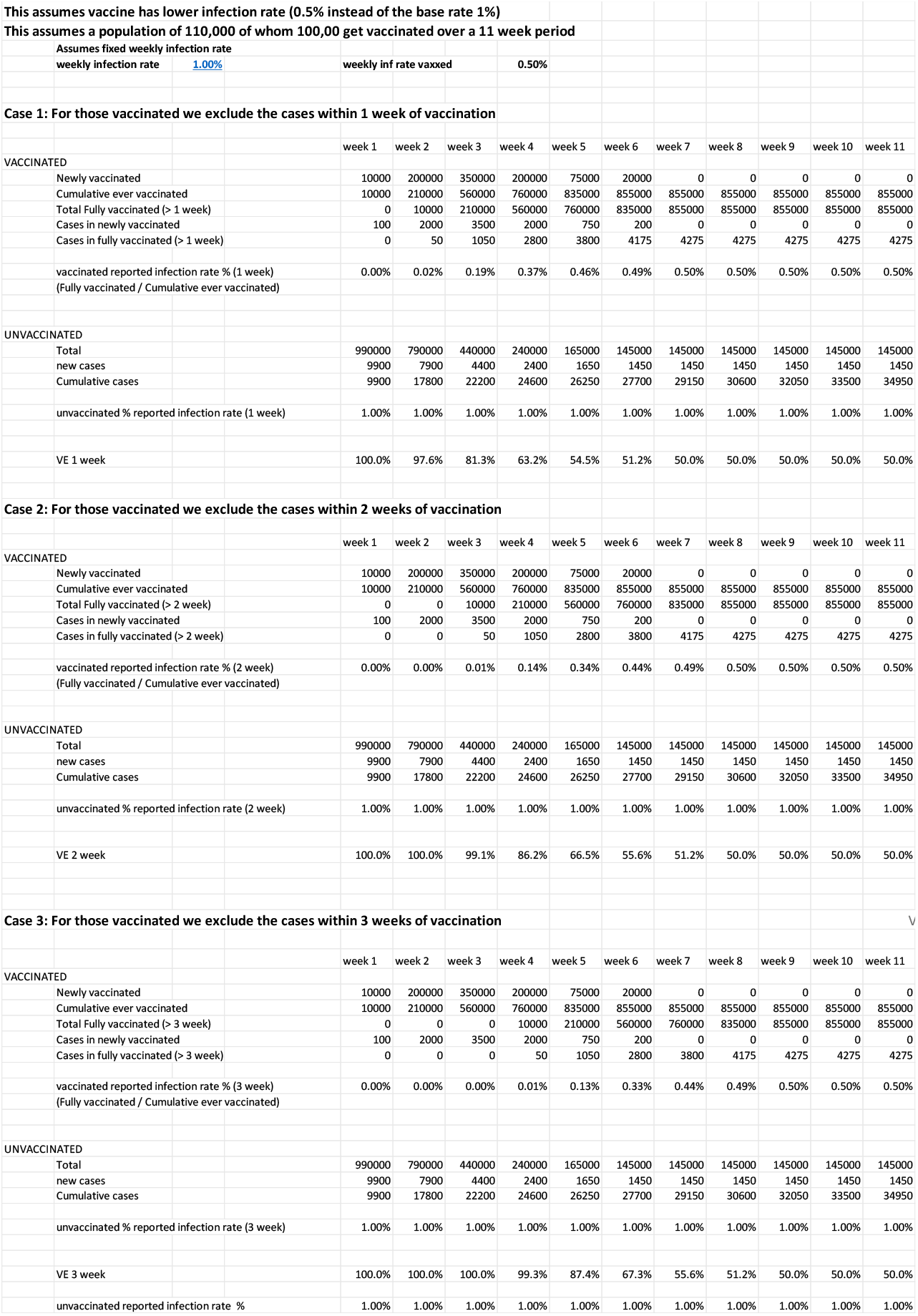

### Scenario H – symmetric exclusion + negative efficacy vaccine

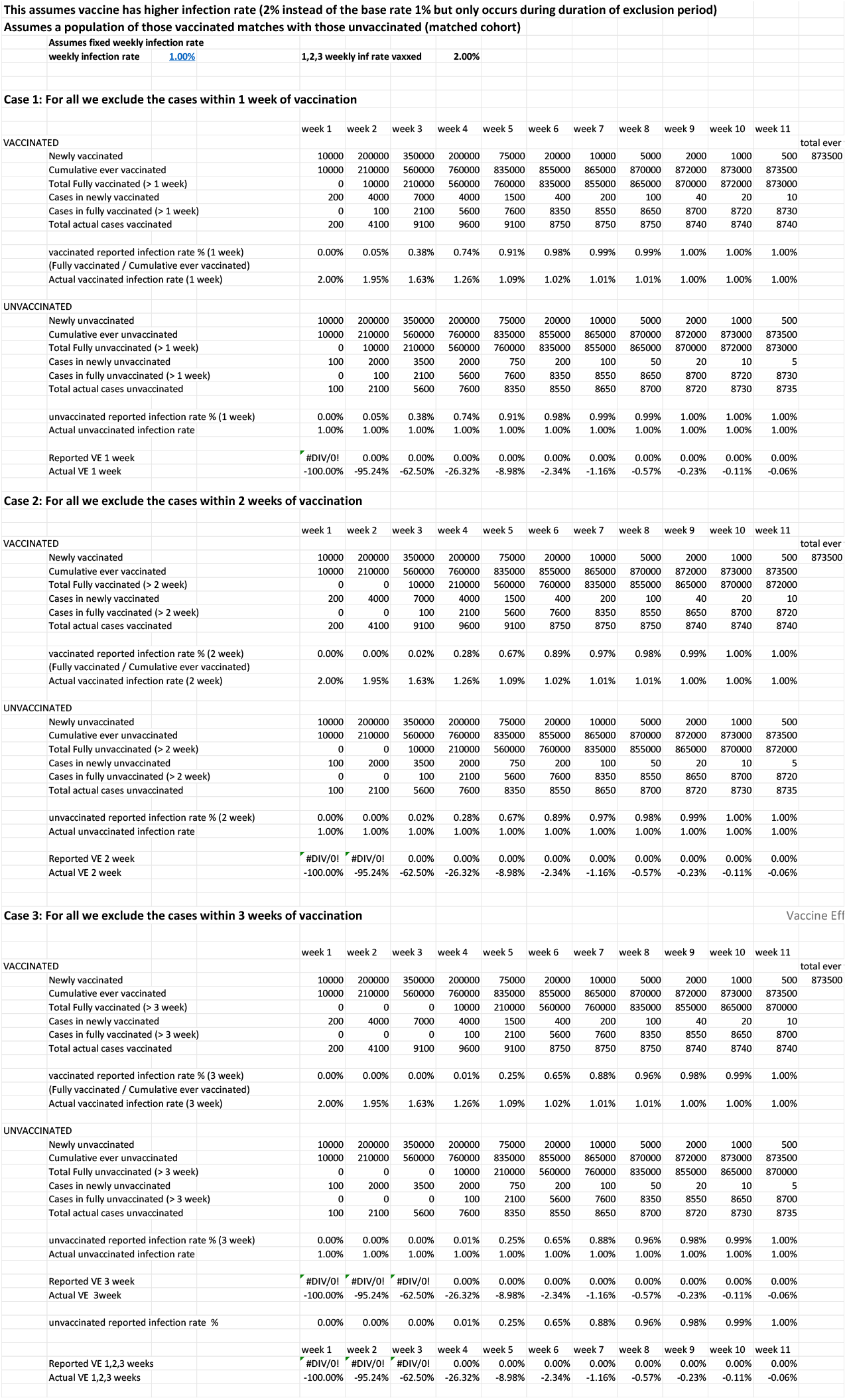

